# Prevalence and risk factors of *Pseudomonas aeruginosa* colonization

**DOI:** 10.1101/2022.05.09.22274871

**Authors:** Nguyen Bao Vy Tran, Quang Minh Truong, Lam Que Anh Nguyen, Ngoc My Huong Nguyen, Quang Hung Tran, Thi Tuyet Phuong Dinh, Vinh Son Hua, Peter A. Lambert, Thi Thu Hoai Nguyen

## Abstract

*Pseudomonas aeruginosa* (*P. aeruginosa*) is one of the most concerning pathogens due to its multidrug resistance. *P. aeruginosa* can be a part of the normal commensal flora of humans but can also cause a wide range of infections. In this study, we investigated the prevalence of commensal *P. aeruginosa* in 609 Vietnamese participants (310 females and 299 males, age range of 2 to 73 years) who had no acute infection or disease symptoms at the time of sample collection. Samples were taken from the throat, naris and outer ears. As a result, 19 were positive with *P. aeruginosa* (3.12%, 95% CI: 0.017-0.045) which came mostly from throat (11/19, 57.89%). Participants with a history of sinusitis were 11.57 times more likely to be colonized with *P. aeruginosa* than participants without a history of sinusitis (OR: 11.57, 95% CI: 4.08-32.76, p-value< 0.0001). Age and gender were not significantly associated with *P. aeruginosa* colonization. The commensal *P. aeruginosa* isolates were tested for biofilm formation, pyocyanin, siderophore, lipase, protease and gelatinase production. Among 16 *P. aeruginosa* isolates used for these tests, 100% (16/16) were positive for biofilm, pyocyanin and siderophores; 93.75% (15/16) isolates were positive for gelatinase and protease; and 50% (8/16) isolates were positive for lipase. There were no differences in the pattern and range of virulence factors of *P. aeruginosa* isolates taken from participants with and without sinusitis history. In summary, *P. aeruginosa* colonized 3.12% of participants, and its presence was clearly associated with sinusitis history. Most commensal *P. aeruginosa* isolates can produce biofilm, pyocyanin, siderophores, gelatinase and protease.

**Author summary:** *P. aeruginosa* is both a common opportunistic pathogen which causes various infections in humans, such as blood, lung, and skin infections and a commensal bacterium which can be found normal human flora. In this study, we showed that the *P. aeruginosa* colonized 3.12% participants and resided mostly in human throat. Interestingly, we found that people with sinusitis history were more likely to be *P. aeruginosa* carriers. On the other hand, age and gender did not significantly affect *P. aeruginosa* colonization. Most tested *P. aeruginosa* isolates expressed various virulence factors, including biofilm, siderophores, pyocyanin, gelatinase, protease, and lipase.

## Introduction

*Pseudomonas aeruginosa (P. aeruginosa)* is a Gram-negative rod-shaped bacterium. It is widely distributed in nature, such as water and soil. *P. aeruginosa* can also be found in the normal flora of humans and is considered as an opportunistic pathogen. Previous data showed that *P. aeruginosa* was found on the skin (0-2%), in the throat (0-3.3%), and in the stool (2.6-24%) [1]. Under continuous pressure from the human immune system, commensal *P. aeruginosa* is capable of transforming into a virulent pathogen [2][3]. In immunosuppressive humans or cystic fibrosis patients, *P. aeruginosa* infection is a major cause of serious health consequences. Unfortunately, infections by *P. aeruginosa* are difficult to treat due to its diverse antibiotic-resistance mechanisms and production of numerous virulence factors that assist its colonization on host cells, such as biofilm formation, proteases, siderophores and exotoxins [4].

Among many virulence factors, biofilm is one of the most important factors which assists *P. aeruginosa* colonization and adaption. Biofilm is a complex structure that contains many planktonic cells sticking together by extracellular polymeric substances [5]. This structure can form on virtually any moist surface such as living tissues, medical devices, water pipes [5][6]. Furthermore, biofilm contributes to the persistence of *P. aeruginosa* infection by protecting this pathogen against host defenses and antimicrobial strategies [7]. Apart from biofilm, extracellular enzymes, such as lipase, protease, gelatinase and hemolysin also contribute to the virulence of *P. aeruginosa* [8]. Besides, pyocyanin, a blue phenazine pigment secreted by *P. aeruginosa* is another potential virulence factor. It damages the host via causing cell respiration dysfunction, calcium homeostasis disruption [9]. In iron-deficient conditions, siderophores play an important role for *P. aeruginosa* persistence as they work as iron chelators to obtain free Fe^3+^ from the environment as well as iron from the iron-binding proteins of humans such as transferrin and lactoferrin [10][11].

Although commensal *P. aeruginosa* can colonize humans without causing any serious problem, *P. aeruginosa* carriers could face the risk of *Pseudomonas* infection. Understanding the properties of commensal *P. aeruginosa* in humans is unexpectedly limited. Thus, this study was undertaken to investigate the prevalence and risk factors of *P. aeruginosa* colonization in a healthy population as well as virulence of the commensal *P. aeruginosa* isolates.

## Results and Discussion

### Prevalence of commensal *Pseudomonas aeruginosa* isolates in Vietnamese population

There were 612 volunteers who signed informed consent and donated samples. Three were excluded due to their declared health condition and 609 participants were finally included in the study (Table 1). The male and female participants were similar, 299 males (49.10%) versus 310 (50.90%) females. Most participants were adults (514, 84.40%); some were younger than 18 years old (65, 10.67%) and few were older than 60 years old (30, 4.93%). It is well-noted that having sinusitis is a common health problem in the population. More than 1/5 of healthy declaring people experienced sinusitis. (Table 1)

**Table 1.** Characteristics of 609 participants in the study

| Characteristics |  | Number<br>Overall (n=609) |
| --- | --- | --- |
| Gender | Male | 299 (49.10%) |
|  | Female | 310 (50.90%) |
|  | 0-17 | 65 (10.67%) |
|  | 18-59 | 514 (84.40%) |
|  | ≥60 | 30 (4.93%) |
| Health status | With sinusitis history | 129 (21.18%) |
|  | Without sinusitis history | 480 (78.82%) |

From 609 participants, 35 *P. aeruginosa*-like isolates were obtained. These isolates were the ones which grew on cetrimide selective media, being Gram-negative rod shape and *orpL*-positive. The *oprL* gene encodes the outer membrane peptidoglycan-associated lipoprotein, which has an important role in the interaction between bacteria and environment, is a common and useful target for *Pseudomonas* detection

In this study, the primer pairs used to detect *oprL*, a common target for *Pseudomonas* detection, were shown to be specific to only *P. aeruginosa* [12], [13], [14]. However, our data showed that the efficiency of this primer pair for detecting commensal *Pseudomonas aeruginosa* was not as expected. Only 20 out of 35 *oprL-* positive isolates (57.14%) were confirmed as *P. aeruginosa* via *16S rRNA* sequencing. This was in agreement with a previous report using the same *oprL*-specific primer pair, in which only 49 out of 70 *oprL-*positive clinical samples (70%) was *P. aeruginosa* [15]. Interestingly, that 100% *oprL-*positive isolates from participants with sinusitis history were confirmed as *P. aeruginosa* indicated that *oprL*-specific PCR could be used to quickly confirm the presence of *P. aeruginosa* in patients with sinusitis history.

Although the *oprL*-specific PCR could not specifically detect *P. aeruginosa*, all 35 *oprL*-positive samples were *Pseudomonas* species, including *P. aeruginosa, P. stutzeri, P. azelaica, P. nitroreducens*. That means the *oprL*-specific PCR could be a potential tool to exclude non-*Pseudomonas* species. Among *oprL-* positive isolates, *P. aeruginosa* (20/35; 57.14%) and *P. stutzeri* (12/35; 34.29%) were 2 dominant *Pseudomonas* commensal isolates. The detail results of *oprL-* positive *Pseudomonas* species that were collected in our study was described in Table 2.

**Table 2.** *oprL-* positive *Pseudomonas* isolates and their colonization sites.

| Species | Total number<br>(n=35) | Throat<br>(n=13) | Naris<br>(n=14) | Outer ears<br>(n=8) |
| --- | --- | --- | --- | --- |
| *P. aeruginosa* | 20 (57.14%) | 11 (84.62%) | 5 (35.71%) | 4 (50%) |
| *P. stutzeri* | 12 (34.29%) | 0 | 9 (64.29%) | 3 (37.50%) |
| *P. azelaica* | 2 (5.71%) | 1 (7.62%) | 0 | 1 (12.50%) |
| *P. nitroreducens* | 1 (2.86%) | 1 (100%) | 0 | 0 |

From our findings, *P. aeruginosa* isolates were likely obtained from the throat (11/20 *P. aeruginosa* isolates), while *P. stutzeri* were rather from the naris area (9/12 *P. stutzeri* isolates). Interestingly, the throat was primarily colonized by *P. aeruginosa* (11/13 isolates in the throat, 84.62%). Only one *P. azelaica*, one *P. nitroreducens* and no *P. stutzeri* were detected in this area.

Among 20 *P. aeruginosa* isolates, two isolates came from one volunteer (from throat and naris) and 18 others came from a single site on a single participant. In summary, 19 out of 609 participants (3.12%, 95%CI: 0.017-0.045) were colonized with *P. aeruginosa*. This was in agreement with previous data showing that *P. aeruginosa* was not commonly detected in healthy people, but under conditions of antibiotic exposure or hospitalization its prevalence, mainly in the throat and stool, was increased [16],[17]. For example, in the case of bronchiectasis, which is a chronic respiratory disease associated with *P. aeruginosa*, the prevalence of *P. aeruginosa* colonization in bronchiectasis patients ranged from 9 to 33% [18]. Besides, Casetta *et al*. also found that 17.5% of pregnant women were colonized by *P. aeruginosa* after more than 48h hospital admission [19].

Importantly, colonization with *P. aeruginosa* is a risk of infection. Gómez-Zorrilla *et al*. reported that 43% of colonized patients in their study developed infection [20]. In hospital setting, the nosocomial infection rate was about 10% in developing countries [21], or about 3-7% in Vietnam [22] and *P. aeruginosa* is one of the leading causes in Vietnam for nosocomial infection [23].

### Relationship between gender and age to *Pseudomonas aeruginosa* colonization

Among 19 commensal *P. aeruginosa* carriers, 63.16% (12/19) were female and 36.84% (7/19) were male. However, there were no associations between gender and *P. aeruginosa* colonization of healthy humans (OR: 1.68, 95%CI: 0.65-4.33, p-value=0.29 and Chi-Square: p-value=0.278). To our knowledge, there is still a lack of information about the effect of gender on *P. aeruginosa* colonization in healthy humans. In contrast to our finding, the study of *P. aeruginosa* colonization from wounds and burns swabs of patients showed the higher rate of *P. aeruginosa* among the male than female samples, 55.6% and 44.4%, respectively [24]. However, the presence of *P. aeruginosa* was associated with a higher morbidity rate in females with respiratory dysfunction, such as cystic fibrosis and bronchiectasis. The estrogen in females could regulate the conversion of *P. aeruginosa* from non-mucoid to mucoid forms, which are believed to have higher virulence [25]. The relationship between gender and *P. aeruginosa* colonization is described in table 3.

**Table 3.** Odds ratio (OR) for gender differences in *P. aeruginosa* carriers. Male was the reference category, hence odds ratio (OR) =1.

| Gender | Number |  | OR | 95%<br>Confidence<br>interval | p-value |
| --- | --- | --- | --- | --- | --- |
|  | *P. aeruginosa*<br>carriers | Non-*P. aeruginosa*<br>carriers |  |  |  |
| **Female**<br>(n=310) | 12 (3.87%) | 298 (96.13%) | 1.68 | 0.65-4.33 | 0.29<br>(>0.05) |
| **Male**<br>(n= 299) | 7 (2.34%) | 292 (97.66%) | 1 |  |  |

Although the group 18-59 years had the highest number of *P. aeruginosa* colonization (84.21%, 16/19 *P. aeruginosa* carriers), age was not a significant factor for *P. aeruginosa* colonization (0-17: 65 participants, 18-59: 514 participants, and ≥ 60: 30 participants) (p-value = 0.448). When compared among each group, the prevalence rate of the age group ≥60 years was highest (6.67%, OR: 6.75, 95%CI: 0.59-77.55, p-value >0.05), the age group 18-59 years was at the second place (3.11%, OR: 2.02, 95% CI: 0.26-15.43, p-value <0.05). The odds ratios (OR) for age differences in *P. aeruginosa* carriers were described in table 4. Currently the association between age and *P. aeruginosa* colonization in healthy humans is unclear, but age >55 years is considered to be an independent predictor for *P. aeruginosa* colonization in bronchiectasis patients [26]. This could be explained by the frequent hospital visiting of the elderly, which increased the chance of contact with *P. aeruginosa* in hospital environments. Another study also described that the incidence of infection by opportunistic pathogens increases significantly in older ages [27] and colonization could be a risk factor for infection [28]. Hence, the presence of opportunistic pathogens could increase when we get older.

**Table 4.** Odds ratio (OR) for age differences in *P. aeruginosa* carriers.

| Age groups, years | Number |  | OR | 95% Confidence interval | p-value |
| --- | --- | --- | --- | --- | --- |
|  | *P. aeruginosa* carriers | Non-*P. aeruginosa* carriers |  |  |  |
| 0-17 (n= 65) | 1 (1.56%) | 64 (98.46%) | 1 |  |  |
| 18-59 (n=514) | 16 (3.11%) | 498 (96.89%) | 2.02 | 0.26-15.52 | 4.96E-02 (<0.05) |
| ≥ 60 (n=30) | 2 (6.67%) | 28 (93.33%) | 6.75 | 0.59-77.55 | 1.25E-01 (>0.05) |

Group 0-17 was the reference category, hence odds ratio (OR) =1.

### Relationship of sinusitis history to *Pseudomonas aeruginosa* colonization

Our data indicated that sinusitis is quite common in the population with 129/ 609 (21.18%) participants having sinusitis history (Table 1). Interestingly, participants with sinusitis history are more likely to be colonized with *P. aeruginosa* (14/129, 10.85%) compared to the ones without sinusitis history (5/480, 1.04%). It is estimated that the participants with sinusitis history have a risk of *P. aeruginosa* colonization 11.57 times higher than that of participants without sinusitis history (OR: 11.57, 95%CI: 4.08-32.76, p-value<0.0001), (Table 5). Our data were somewhat in agreement with previous studies. For example, Niederman *et al*. indicated that the upper and lower respiratory tract was not colonized by Gram-negative bacteria under normal health conditions, but these sites could be harbored by these pathogens when illness developed [29]. In the case of cystic fibrosis patients, Shapiro *et al*. found that *P. aeruginosa* was detected in 38% of sinusitis-cystic fibrosis patients [30] while Kasper Aanæs *et al*., reported that the presence of *P. aeruginosa* in sinuses was associated with lung infection in cystic fibrosis patients [31]. However, data were also quite controversial as in some studies, *P. aeruginosa* was not a common pathogen in bacterial flora of chronic sinusitis patients, only present for 1% to 5% of cases [32],[33],[34].

**Table 5.**
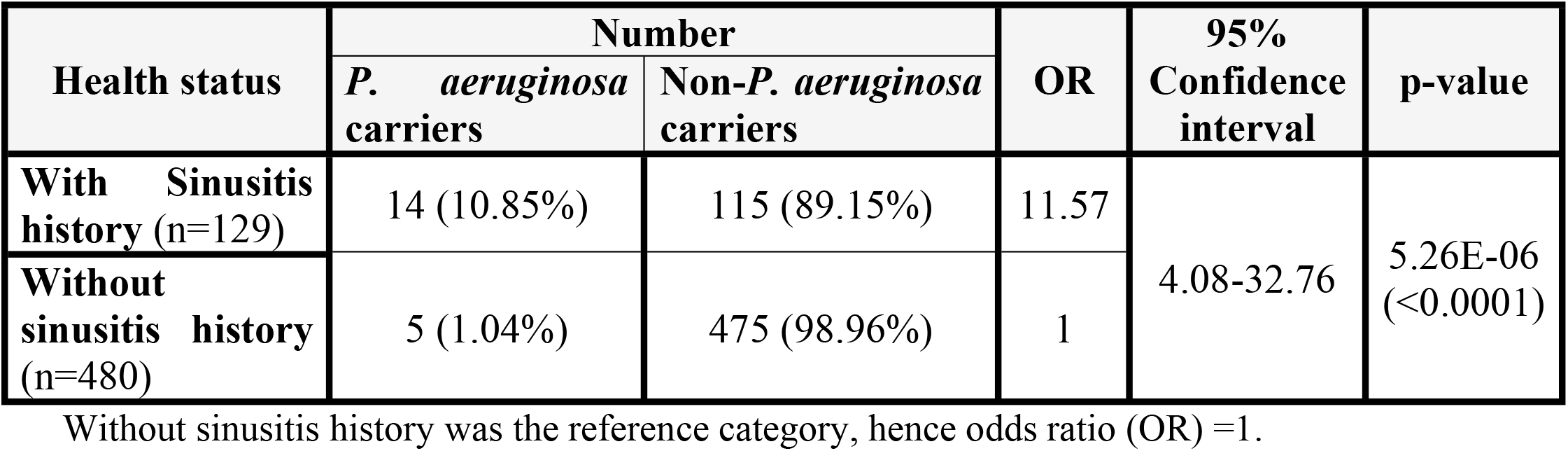
Relationship of sinusitis history and *P. aeruginosa* carriage.

### Commensal *Pseudomonas aeruginosa* virulence

In this study, 16 out of 20 *P. aeruginosa* isolates were used for testing the production of key virulence factors, including biofilm, pyocyanin, siderophores, lipase, protease, and gelatinase [Table S1]. 100% (16/16 *P. aeruginosa* isolates) isolates were positive for biofilm, pyocyanin, siderophores and hemolysin; 93.75% (15/16) isolates were positive for gelatinase, protease; and 50% (8/16) isolates were positive for lipase (Table 6).

**Table 6.** Percentage of positive isolates and average virulence value of biofilm, pyocyanin, siderophores, lipase, protease and gelatinase assays.

| Virulence factors | Positive | Average virulence value ± |
| --- | --- | --- |

|  | percentage (%) | standard deviation |  |
| --- | --- | --- | --- |
|  |  | Commensal *P. aeruginosa* isolates | *P. aeruginosa* ATCC 9027 |
| Biofilm (OD550nm) | 100% | 0.11 ± 0.06 | 0.72 ± 0.10 |
| Pyocyanin (ug/mL) | 100% | 0.61 ± 0.40 | 0.60 ± 0.23 |
| Siderophores (mm) | 100% | 1.31 ± 0.34 | 1.33 ± 0.29 |
| Lipase (mm) | 50% | 0.18 ± 0.04 | 0.17 ± 0.03 |
| Protease (mm) | 93.75% | 0.28 ± 0.09 | 0.17 ± 0.03 |
| Gelatinase (mm) | 93.75% | 0.29 ± 0.06 | 0.22 ± 0.03 |

All tested isolates showed the ability to produce biofilm (16/16). Among them, there were 25% (4/16) of weak, 56.25% (9/16) of moderate, and 18.75% (3/16) of strong biofilm producers.

Similarly, all tested commensal *P. aeruginosa* isolates had the ability to secrete phenazine pyocyanin. The concentration ranged from 0.05 µg/mL to 1.28 µg/mL with the average was 0.61 ± 0.40 (µg/mL) (mean ± standard deviation).

Following CAS blue agar method, all tested commensal isolates could synthesize and secret siderophores to obtain iron in the environment. However, there was only one isolate indicated as a moderate siderophore producer (sample 464.3), all others were weak siderophore-producing isolates (93.75%, 15/16). The size of the pale yellow halo zone ranged from 0.75 mm to 2.17 mm. The average of the halo zone sizes was 1.31 ± 0.34 (mm). While 93.75% of the tested *P. aeruginosa* produced protease and gelatinase, their average enzymatic activity was 0.28 ± 0.09 (mm) and 0.29 ± 0.06 (mm), respectively. Only 50% of tested isolates positive for lipase, which the average halo size was 0.18 ± 0.04 (mm) (Table 6).

Although sinusitis was a risk factor for *P. aeruginosa* colonization, there were no significant differences in virulence between *P. aeruginosa* isolates from carriers with and without sinusitis history (Table 7).

**Table 7.** The virulence of commensal *P. aeruginosa* isolates isolated from participants with and without sinusitis history. Average ± standard deviation.

|  | With sinusitis history | Without sinusitis history | p-value (ANOVA) |
| --- | --- | --- | --- |
| **Biofilm** (OD<sub>550nm</sub>) | 0.13±0.06 | 0.20±0.3 | 0.39 |
| **Pyocyanin** (ug/ml) | 0.57±0.41 | 0.71±0.36 | 0.52 |
| **Siderophores** (mm) | 1.35±0.39 | 1.22±0.09 | 0.48 |
| **Lipase** (mm) | 0.14±0.04 | 0.17±0.03 | 0.27 |
| **Gelatinase** (mm) | 0.28±0.1 | 0.24±0.03 | 0.40 |
| **Protease** (mm) | 0.27±0.11 | 0.22±0.08 | 0.41 |

In this study, there was a positive correlation between biofilm formation and siderophores, protease secretion in commensal *P. aeruginosa* (p-value < 0.05). This is consistent previous studies indicating that siderophores were considered as a signal for biofilm development [35] and biofilm formation decreased when the related siderophore genes were deleted [36].

Furthermore, the commensal *P. aeruginosa* isolates showed significant weaker production of the virulence factors in compared to clinical isolates. These included biofilm production (0.11 ± 0.06, compared to 1.39 ± 0.17, p-value < 0.05), pyocyanin (0.61 ± 0.40, compared to 1.92 ± 0.56 ug/mL, p-value< 0.05), siderophores (1.31 ± 0.34, compared to 5.02 ± 0.42 mm, p-value < 0.05), lipase (0.18 ± 0.04, compared to 1.23 ± 0.69, p-value< 0.05), protease (0.28 ± 0.09, compared to 2.71 ± 1.33, p-value <0.05), and gelatinase (0.29 ± 0.06, compared to 2.45 ± 1.12, p< 0.05) (Table S6, S7). This suggested that commensal *P. aeruginosa* isolates significantly produced less virulence factors than clinical isolates.

## Materials and Methods

### Commensal *P. aeruginosa* isolation

From 2018 to 2020, throat, naris, and outer ear swab samples of Vietnamese living in the Southeast area were collected and cultured on *Pseudomonas* selective Cetrimide media. All volunteers or volunteers’ guardians gave their signed informed consent. Only people with no declared current acute infection or disease symptoms were included in the study. Colonies obtained on Cetrimide agar (HiMedia, India) were further characterized using Gram-staining, polymerase chain reaction (PCR) with *oprL* primers as a screening step, and *16S rRNA* sequencing for confirmation. For further experiments, all the *P. aeruginosa* isolates were stored in Luria Bertani (LB, HiMedia, India) broth containing 30% glycerol at -80°C.

### *P. aeruginosa* identification

#### DNA extraction

The organic phenol-chloroform method was applied to extract DNA from a single colony of each isolate. The quality of extracted DNA was supposed to be good if the ratio 260/280 was in the range of 1.8 to 2.0. For long-term storage, DNA was stored in Tris-EDTA at -20°C.

#### *oprL*-specific polymerase chain reaction and *16S rRNA* sequencing

The specific *oprL* primers were used to primary detect *P. aeruginosa* (forward: 5’-ATGGAAATGCTGAAATTCGGC-3’ and reverse: 5’-CTTCTTCAGCTCGACGCGACG-3’) [13]. Each PCR reaction had a total volume of 25 µL containing 12.5 µL master mix (iStandard iVAPCR Master Mix, VietA Corp, Vietnam), 8.5 µL free nucleotide water, 0.4 uM of each primer (PHUSA Biochem Oligo, Vietnam) and 2 µL extracted DNA. The PCR program had an initial denaturation of 2 minutes at 95°C, followed by 30 cycles of denaturation at 95°C for 2 minutes, annealing at 56°C for 30 seconds, and extension at 72°C for 1 minute, finally incubation for 5 minutes at 72°C [13].

All positive isolates for *oprL* were sent to NAM KHOA Biotek Company for *16S rRNA* sequencing. The confirmed *P. aeruginosa* isolates were used for further experiments.

### Virulence testing

Biofilm, pyocyanin, siderophores, lipase, protease, and gelatinase of the isolated *P. aeruginosa* isolates were analyzed. Before testing, each sample was streaked on selective Cetrimide agar. Single colonies of each isolate were inoculated in 5 ml of LB broth overnight at 37°C to achieve optimal density (OD_620nm_) approximately 0.08-0.1 to make sure every isolate had the same bacterial concentration before virulence tests. Each virulence test was performed in triplicate.

#### Biofilm

Biofilm production of commensal *P. aeruginosa* isolates was tested by the crystal violet staining method. The optimal density (OD) was measured by ELISA microplate reader at 550 nm [37]. Commensal isolates were classified into negative, weak, moderate and strong biofilm producers by following this category: OD ≤ ODc (negative), ODc < OD ≤ ×2ODc (weak), 2×ODc < OD ≤ 4×ODc (moderate) and OD > 4×ODc (strong), in which ODc was the cut-off value (ODc = OD negative control + 3 × its standard deviation) [38]. *P. aeruginosa* ATCC 9027 was used as a positive control for the test.

#### Pyocyanin

Pyocyanin was extracted and measured using the chloroform method. Glycerol-alanine (Gly-Ala) broth medium was used to maximize the yield of pyocyanin production in commensal isolates [39]. The absorbance was measured by using ELISA microplate reader at 520 nm. The concentration of pyocyanin (μg/mL) was estimated by multiplying the optical density (OD) at 520 nm by 17.072 [40][41]. *P. aeruginosa* ATCC 9027 was used as a positive control for the test.

#### Siderophores, lipase, protease, and gelatinase

These virulence factors were detected using agar-based methods. The *P. aeruginosa* cultures were spotted on CAS blue agar, tributyrin agar (1% tributyrin), skim milk (3% skim milk, Brain Heart Infusion (BHI) medium), and gelatin agar (8% gelatin, BHI (NamKhoa Biotek, Vietnam), which were specific to siderophores, lipase, protease, and gelatinase, respectively. All the cultures were incubated at 37°C for 24 hours, except sheep blood agar plates for 72 hours. For the gelatinase test, (NH_4_)_2_SO_4_ solution was added before result observation [42][43][44]. The size of the halo zone around the spot corresponds to the virulence factor-producing ability or enzymatic activity of commensal *P. aeruginosa* isolates. The enzymatic activity (EA) was estimated using the formula: EA = (D-d)/2, where D was the diameter of clear zone (mm) and d was the diameter of a colony (mm) (Vermelho et al. 1996). It was categorized as negative (0 mm), weak (<2mm), moderate (2-4 mm), and strong (>4 mm) activity [45]. *P. aeruginosa* ATCC 9027 was used as a positive control for all the tests, while *Staphylococcus aureus (S. aureus)* was a negative control for siderophores.

### Data analysis

Each experiment was performed in triplicate. IBM® SPSS® Statistics 20.0 (IBM, USA) was used to analyze the data. Chi-square and Fisher’s Exact tests were used to determine the risk factors for *P. aeruginosa* colonization. The Mann-Whitney U test was used to indicate the significance between commensal and clinical virulence data. For the relationship between virulence factors of commensal *P. aeruginosa*, linear regression was performed. In addition, an ANOVA analysis was used to show the association between sinusitis history and commensal *P. aeruginosa* virulence. P-value was set to be <0.05.

### Ethical approval

This study was approved by the Ethics Committee of Vietnam National University of Ho Chi Minh City (Date 06/06/2019 /No 1007/DHQG-KHCN). Informed consent was sent and explained to all volunteers before any sample collecting process.

## Conclusion

The prevalence of commensal *P. aeruginosa* in the Vietnamese population was 3.12%. Sinusitis could be a potential factor contributing to the colonization of *P. aeruginosa*. The commensal *P. aeruginosa* isolates can produce multiple virulence factors including biofilm, pyocyanin, siderophores, lipase, protease, gelatinase and lipase. However, the virulence values of these commensal *P. aeruginosa* isolates were significantly lower than the clinical *P. aeruginosa* virulence. There was no correlation between commensal *P. aeruginosa* virulence and sinusitis history.

## Data Availability

All data produced in the present study are available upon reasonable request to the authors

## Acknowledgements

This research is funded by Vietnam National University of Ho Chi Minh City under grant number: C2020-28-03.

## Notes

### Competing Interest Statement

The authors have declared no competing interest.

### Funding Statement

This study was funded by Vietnam National University of Ho Chi Minh City under grant number: C2020-28-03.

### Author Declarations

Ethics committee of Vietnam National University of Ho Chi Minh City gave ethical approval for this work

